# Environmental risk factors for gastric cancer: a protocol for umbrella review of meta-analyses

**DOI:** 10.1101/2020.09.20.20196345

**Authors:** Min Seo Kim, Seungwon Kim, Sungsoo Park

## Abstract

Gastric cancer is the fifth most commonly diagnosed cancer globally, accounting for 5.7% of the new cancer cases in 2018[1]. Gastric cancer remains an important global concern because it is the third leading cause of cancer death[1] and its increasing incidence[2]. Therefore, it is crucial to develop prevention strategies for gastric cancer. Helicobacter pylori infection, a WHO class I carcinogen, is a significant risk factor of gastric cancer; however, only 0.5% of infected individuals would develop gastric cancer[3]. This data suggests that there are other additional risk factors for gastric cancer. A large body of research has been performed to investigate the association between various environmental factors and gastric cancer, including dietary factors, tobacco smoking, and alcohol drinking. However, the precision and strength of these existing studies need to be clarified. Umbrella review is one of the most useful tools for a clear understanding of a broad topic area. It provides a comprehensive overall picture of findings for specific questions based on high-level evidence of published systemic reviews and meta-analyses[4]. Thus, this study aimed to conduct an umbrella review of systemic reviews and meta-analyses that investigated gastric cancer risk factors.

## METHODS

### Eligibility Criteria

#### Condition or domain being studied

Our domain of study will be the incidence of gastric cancer

#### Participants/population

Patients who have not been diagnosed with GC prior to initiation of the study or have recorded disease-free status will be included for analysis.

#### Interventions/exposures

Diverse environmental risk factors such as smoking, alcohol, coffee, green tea, grain, socioeconomic status, education, statin or PPI intake, spicy and chili consumption, HTLV-1, EBV, H. pylori, asbestosis, carotenoids, cement, and all other factors that have been investigated for the association to GC are subject to our investigation.

#### Comparator/control

Comparator will be non-exposed or low-exposed group.

#### Types of Studies (inclusion and exclusion criteria)

Studies of the following forms will be included:

- meta-analyses of observational studies.
- systematic reviews presenting quantitative synthesis such as forest plots.
- studies investigated on the general population.

Studies of the following forms will be excluded:

- narrative review.
- meta-analyses with insufficient data for quantitative synthesis or re-analysis.
- outdated meta-analyses (more than 5 years gap from the latest meta-analysis for same topic)
- systematic reviews without meta-analyses for main outcomes.
- studies focusing on polymorphism or genetic instruments.
- studies presenting results in prevalence rather than incidence of gastric cancer.
- studies published in languages other than English and thus cannot examine the study to its fullest extent.
- studies exclusively investigating specific populations and thus unable to extrapolate the results to general populations (e.g., exclusively with child/adolescent, elderly, black, Asian, etc.)
- studies exploring treatment responses or prognostic/survival outcomes of gastric cancer.
- studies involving animal experiments or in vitro results.

### Primary Outcomes

The main outcomes will be the incidence/occurrence of gastric cancer. Measures of the effect of risk factors on GC can be presented as metrics such as odds ratio (OR), relative risk (RR), hazard ratio (HR), standardized incidence ratio (SIR), and standardized mortality ratio (SMR). We will preserve metrics as reported in the original meta-analyses and avoid rough conversion of effect size measures.

### Search Methods for Identification of Studies

#### Electronic Database Search

We will search on PubMed, Embase, and The Cochrane Library (Cochrane database of systematic reviews) from inception to September 2020. Two researchers (MS Kim and S Kim) will independently search for systematic reviews and meta-analyses investigating diverse environmental risk factors known to affect the developing of gastric cancer.

##### PUBMED

(gastric cancer* OR (cancers,gastric) OR “stomach cancer*” OR “gastric neoplasm*” OR (neoplasms, gastric) OR “stomach neoplasm*” OR “gastric malignanc*” OR “stomach malignanc*” OR “gastric tumor*” OR “stomach tumor*” OR “Stomach Neoplasms”[Mesh]) AND (meta-analysis[ptyp] OR meta[ti] OR meta-analy*[ti] OR pooled[ti] OR mendelian[tiab] OR “Meta-Analysis”[publication type]) AND (risk*[tiab] OR incidence*[tiab] OR association*[tiab] OR associat*[tiab] OR “Risk”[Mesh] OR “Incidence”[Mesh] OR “Association”[Mesh]) NOT gastrectom*[ti] NOT surviv*[ti] NOT prognos*[ti] NOT protocol*[ti] NOT comment*[ti] NOT polymorphism*[ti] NOT kid*[ti] NOT child*[ti] NOT adolesc*[ti] NOT pediatric*[ti]

##### EMBASE

(stomac*:ti,ab,kw OR gastri*:ti,ab,kw) AND (cancer*:ti,ab,kw OR tumor*:ti,ab,kw OR neoplasm*:ti,ab,kw) AND (meta:ti OR ‘meta analy*’:ti OR pooled:ti) AND (risk*:ti,ab,kw OR incidenc*:ti,ab,kw OR associat*:ti,ab,kw) NOT (gastrectom*:ti OR surviv*:ti OR prognos*:ti OR protocol*:ti OR comment*:ti OR polymorphism*:ti OR kid*:ti OR child*:ti OR adolesc*:ti OR pediatric*:ti) NOT (‘conference abstract’:it OR ‘conference paper’:it OR ‘conference review’:it OR editorial:it OR note:it OR letter:it OR ‘short survey’:it)

##### CDSR

(stomach* OR gastri*) AND (cancer* OR neoplasm* OR tumor*)

### Data Collection and Analysis

This part is similar to the description in our previous umbrella review protocols[5, 6] as our working frame and basic study design for umbrella review do not differ.

#### Data extraction (selection and coding)

Two researchers (MS Kim and S Kim) will independently search the literature using aforementioned search strategy. Titles, abstracts, and full text of individual study will be reviewed for inclusion and any duplicate will be check for removal. Any discrepancy between authors during the process will be resolved through a decision-making by a third party (S Park, corresponding author). The study selection process will be recorded using PRISMA flowchart.

A predefined data extraction table will be used to extract data and summarize each study. Following details will be obtained: population characteristics, exposure, comparison, measure/index, publication year, number of included studies in a meta-analysis, number of events and cohorts, reported summary effect size, heterogeneity I^2^, and outcome of interest.

We will preferentially use pooled effect sizes of studies that present all individual studies (e.g., forest plot) rather than rely on summary pooled effect sizes derived from studies without much information on individual studies.

#### Risk of Bias (quality) assessment

We will assess the study quality of included systematic review and meta-analysis using the validated AMSTAR 2 (A Measurement Tool to Assess Systematic Reviews 2) instrument. Assessing the risk of bias of individual studies included in the meta-analyses is beyond our scope and will not be conducted; this work is subject to each author.

We will evaluated the certainty of evidence for each main/primary outcome using GRADE (Grading of Recommendations Assessment, Development and Evaluation) approach, as has been done in numerous previous umbrella reviews[7-10]. Small study effect will be assessed with Egger’s test of funnel plot asymmetry.

#### Strategy for data synthesis

We will replicate the meta-analyses in our analytic framework and re-analyzed the data to uncover the non-explicit details such as heterogeneity (I^2^), Egger’s p values, prediction interval, and effect size in the fixed-effect model; this information will be used in appraising the quality of evidence for each outcome. We will re-analyze each meta-analysis under both fixed and random effects model, adhere to the workflow for pairwise meta-analyses described elsewhere[11], and effect sizes and 95% confidential intervals (CI) will be pooled. We will preserve the metrics reported in the original meta-analyses (RR, OR, HR, etc). According to GRADE framework, the results of the main outcomes (but not subgroup analyses) will be used to construct an evidence map. Software R and its packages will be used for the analysis.

#### Subgroup and Sensitivity Analyses

Subgroup/sensitivity analyses will be performed by following factors when applicable:

- sex (men and women)
- location (e.g. Eastern vs Western)
- anatomical location of GC (e.g. cardiac GC and non-cardiac GC)
- histological type of GC (e.g. diffuse type, intestinal type, etc)

In many cases, subgroup analyses presented by original meta-analyses do not offer a complete set of data needed to calculate certainty of the evidence (e.g., prediction interval, heterogeneity, Eggar’s test), and such low density of information may introduce bias. Therefore, outcomes from subgroup/sensitivity analyses are not subject to bias and will not be included in the evidence map/evidence level stratification.

## Data Availability

Contact to

## ACKNOWLEDGEMENTS

None.

## AUTHOR STATEMENT

None

## CONFLICTS OF INTEREST

The authors declare no potential conflicts of interest.

## AUTHOR APPROVAL

All authors have been seen and approved the protocol

## Notes

### Competing Interest Statement

The authors have declared no competing interest.

